# Socio-educational Impact and Psychological Distress of Medical Students amid the COVID-19 Pandemic: A Japanese Cross-Sectional Survey

**DOI:** 10.1101/2020.10.22.20216572

**Authors:** Yoshito Nishimura, Kanako Ochi, Kazuki Tokumasu, Mikako Obika, Hideharu Hagiya, Hitomi Kataoka, Fumio Otsuka

## Abstract

The COVID-19 pandemic has negatively affected medical education. However, few data are available about medical students’ distress during the pandemic. This study aimed to provide details on how medical students had been affected by the pandemic. In this cross-sectional study, 717 medical students participated in the web-based survey. The questions included how their mental status had changed before and after the Japanese nationwide state of emergency (SOE). 65.9% (473/717) participated in the study. 29.8% (141/473) reported concerns about the shift toward online education, mostly because they thought online education could have been ineffective compared with in-person learning. Participants’ subjective mental health status significantly worsened after the SOE was lifted (p <.001). Those who had concerns about a shift toward online education had higher odds of having generalized anxiety and being depressed (OR 1.97, 95% CI 1.19 – 3.28), as did those who requested food aid and mental health care resources (OR 1.99, 95% CI 1.16 – 3.44; OR 3.56, 95% CI 2.07 – 6.15, respectively). Given our findings, the sudden shift to online education might have overwhelmed medical students. Thus, we recommend educators to inform learners that online learning is non-inferior to in-person learning, which could attenuate potential depression and anxiety.

## Introduction

The coronavirus disease 2019 (COVID-19) global pandemic has drastically changed our lives, with more than eight million cases and 400,000 deaths reported globally as of June 18, 2020, according to the World Health Organization statistics(1). In Japan, 17,668 cases have been confirmed as of June 18, 2020, with an explosion in the number of cases in early April(2). While Japan seems to be bringing the outbreak under control through active cluster tracing, restriction of mass gatherings, and advocating universal masking and hand hygiene, there were no simple solutions to this issue(3). After the surge in COVID-19 cases, on April 16, 2020, the Government of Japan (GoJ) declared a state of emergency (SOE) in all 47 prefectures, which lasted until May 25, 2020(4). In Okayama, a prefecture in the western part of mainland Japan(5) with approximately 1.9 million people (ranked 20/47 among the 47 prefectures), only 25 confirmed cases had been reported by the end of May 2020. Behind the scenes of successful COVID-19 mitigation, however, those in education-related jobs and medical students struggled with the rapid change in the socio-educational system.

Although the SOE was lifted on May 14 in Okayama based on the low incidence of COVID-19, even before the SOE, from March 25, Okayama University School of Medicine (OUSM), one of the largest national universities in Japan, requested its students to stay home to prevent the possible community spread of COVID-19 within the medical school and hospital. Until the stay-home request was lifted on May 22, medical students were required to cope with this sudden change in their lifestyle, and shifted to online education amid the fear and considerable uncertainty surrounding COVID-19. In the early 2000s, the severe acute respiratory syndrome coronavirus (SARS-CoV) outbreak had a devastating impact on academic education, including sudden curriculum changes and rapid integration of information technology(6–8). Similarly, the current COVID-19 pandemic has provoked significant turmoil in society. In particular, mental health problems due to the pandemic have drawn attention worldwide as studies have suggested the need for mental health care interventions during the outbreak(9–14). Medical students, who essentially need clinical exposure, may have been impacted even further by the pandemic.

In Japan, medical schools have different admission practices and curricula than in the United States. Students typically enter medical school immediately after high school graduation, often at 18 years old, and they go through six years of medical education before graduation. Despite the differences in education system, the fundamental philosophy of academic medicine is the same; to provide quality educational experience. To date, few published studies have investigated the status of medical students’ living environment, socio-educational, and mental health status during the COVID-19 pandemic. To address this, we conducted a total population survey of OUSM medical students to comprehensively clarify what they require and how they have been mentally affected by COVID-19.

## Methods

### Study Design, setting, and participants

We performed a cross-sectional study that employed an anonymous, self-administered voluntary web-based survey. Participants comprised medical students in all years of study at OUSM. We used purposive sampling to conduct a total population sample for the 717 medical students who belonged to the OUSM as of April 1, 2020 (the first day of the academic year in Japan). The participants’ consent was implied by their completion of the survey.

The survey was developed through consultation with a medical education expert panel at OUSM and piloting, and administered with Qualtrics (Qualtrics International Inc., Provo, Utah), a web-based survey platform. We provided survey instructions and instruments in Japanese. We distributed survey links to the students using OUSM official mailing lists. All participants were invited to complete the survey within one week (June 8–14, 2020, in Japan Standard Time). No financial incentives were provided for their participation in the survey. In the survey, we included entries on demographics (age, gender, education before entering medical school, employment status on date of response, changes in employment status due to the COVID-19 pandemic, marital status, living environment, household size, and comorbidities) as well as COVID-19 related items (e.g., “Chance of contracting COVID-19 during the current pandemic”), self-learning associated activities (e.g., Average time of self-learning/day), validated depression and anxiety scale instruments, and financial situations. To protect participants’ anonymity as much as possible, respondents were not prompted to enter their year of study.

### Measurements

#### COVID-19-related questions

To evaluate the extent of participants’ concern and preparedness for COVID-19, they were asked: “What do you think the chances are that you will contract COVID-19 during the current pandemic?”; “What is your degree of concern about the health status of your family?”; “Do you think you have enough information about the symptoms of COVID-19?”; “Do you think you have enough information about prevention and treatment of COVID-19?”; and “Do you feel worried about COVID-19?” To further assess students’ concerns, the participants were prompted to answer “I am concerned because my future career formation may be negatively affected due to the COVID-19 pandemic”; “I am concerned because the COVID-19 pandemic may attenuate our relationship to teachers”; “I am concerned because of the disruption to ongoing research or extracurricular activities”; and “I am concerned about the shift toward online education.” All questions were evaluated on a 5-point Likert scale except for the item on the health status of the family (3-point Likert scale). Those who answered “Very concerned” or “Concerned” in the entry on concern about the shift toward online education were prompted to provide reasons. To describe participants’ needs, they were asked to note the types of support they wish to receive from the university if there is a resurgence of COVID-19. These responses were mandatory.

#### Self-learning and related activity

For subjective mental health status and the average time per day that participants stayed at home, read books, played video games, and learned by themselves, respondents were prompted to answer “How many hours a day did you… / stay at home / read books / play video games / self-learn?” before the SOE order (April 16, 2020) or during the last two weeks (with the base date of when participants completed the survey).

#### Depression and anxiety disorders

We assessed the presence of depression using the 9-item Patient Health Questionnaire (PHQ-9), a common screening tool for mood disorders. We used the validated Japanese translation of the scale(15). The total scores of PHQ-9 range from 0 to 27 and we defined scores of 10 or more as having “depression.” We screened for anxiety disorders using the Japanese version of the 7-item Generalized Anxiety Disorder Scale (GAD-7), which was validated in 2010(16). The score ranges from 0 to 21, and we defined scores of 10 or more as having “anxiety.” Both instruments ask respondents about their mental health status during the last two weeks.

#### Financial situation

In Japan, university students typically live on a monthly allowance from parents. According to the latest statistics by the National Federation of University Co-operative Associations, students receive 72,810 JPY (approximately 680 USD) a month on average(17). Participants were prompted to give their monthly allowance from the following options: “None,” “0,” “< 30,000 JPY (280 USD),” “30,000 – 49,999 JPY (280 – 467 USD),” “50,000 – 69,999 JPY (467 – 654 USD),” “70,000 – 99,999 JPY (654 – 935 USD),” and “≥ 100,000 JPY (≥ 935 USD).” Respondents were also asked to answer if they were on scholarship or student loan.

### Statistical analysis

We analyzed the data using JMP version 13.1.0 (SAS Institute Inc., Cary, North Carolina). We used the Wilcoxon signed-rank test to examine differences in the time participants spent on self-learning related activities based on non-normal distribution. For associations between categorical variables containing small sample sizes, we employed Fisher’s exact test. To examine the predictive factors of categorical dependent variables, we used univariate logistic regression analyses. The threshold for significance was defined as *p* < 0.05.

### Ethical approval

This study protocol was approved by the institutional review board of Okayama University Hospital (reference number 2006-029, approved on June 5, 2020).

### Data Availability Statement

The datasets generated and analysed during the current study are available from the corresponding author on reasonable request.

## Results

Response rates to the survey were 473 out of 717 (65.9%) OUSM students in all six years combined. Participants’ demographic characteristics are summarized in **Table 1**. Of note, 250 (52.9%) respondents reported that they were engaged in part-time work, while 44 (9.3%) reported having resigned or been fired due to the COVID-19 pandemic. Eight (1.7%) and 6 (1.3%) noted that they had a past medical history of anxiety disorders and depression, respectively.

**Table 1.**
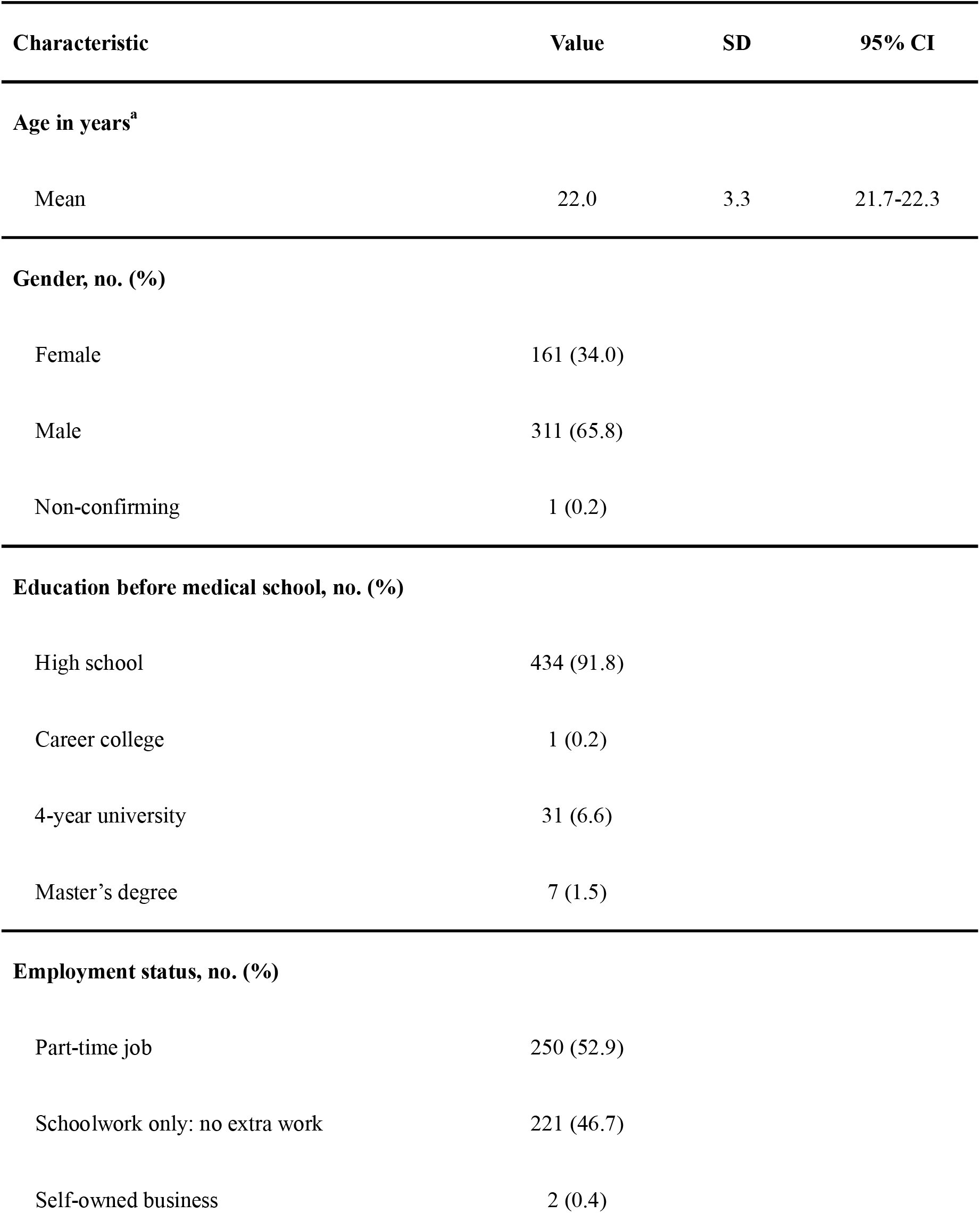
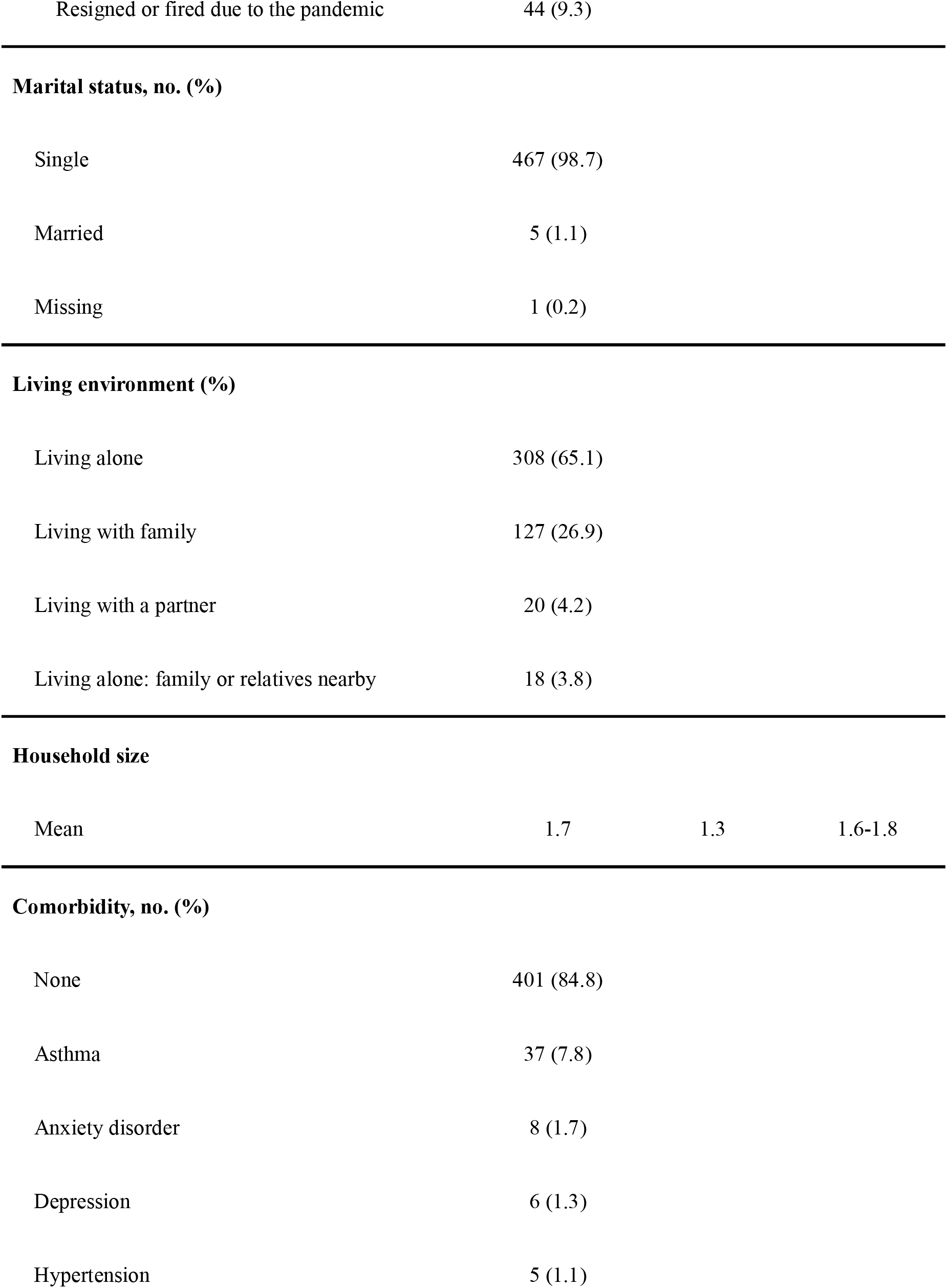
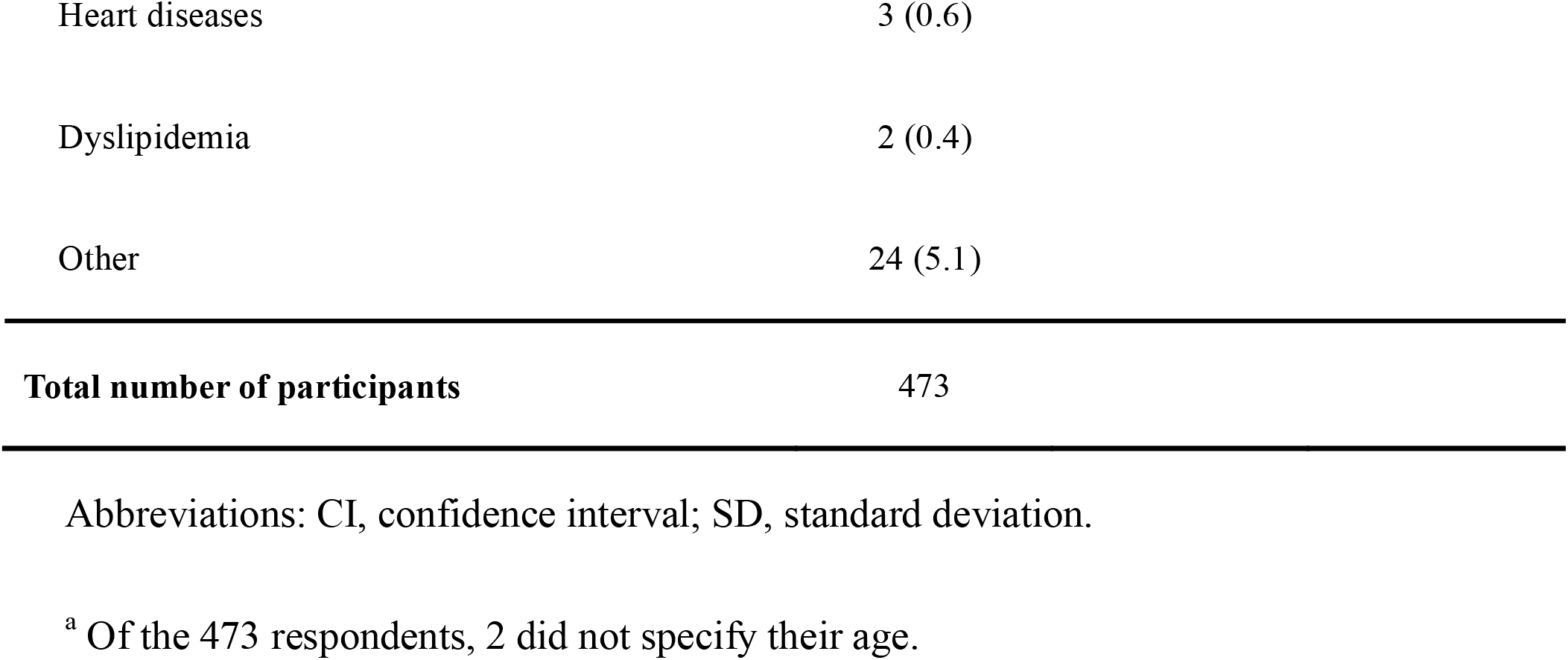
Demographic Characteristics of the Study Participants Comprising 473 Japanese Medical Students.

| Characteristic | Value | SD | 95% CI |
| --- | --- | --- | --- |
| **Age in years<sup>a</sup>** |  |  |  |
| Mean | 22.0 | 3.3 | 21.7-22.3 |
| **Gender, no. (%)** |  |  |  |
| Female | 161 (34.0) |  |  |
| Male | 311 (65.8) |  |  |
| Non-confirming | 1 (0.2) |  |  |
| **Education before medical school, no. (%)** |  |  |  |
| High school | 434 (91.8) |  |  |
| Career college | 1 (0.2) |  |  |
| 4-year university | 31 (6.6) |  |  |
| Master's degree | 7 (1.5) |  |  |
| **Employment status, no. (%)** |  |  |  |
| Part-time job | 250 (52.9) |  |  |
| Schoolwork only: no extra work | 221 (46.7) |  |  |
| Self-owned business | 2 (0.4) |  |  |
| Resigned or fired due to the pandemic | 44 (9.3) |  |  |
| **Marital status, no. (%)** |  |  |  |
| Single | 467 (98.7) |  |  |
| Married | 5 (1.1) |  |  |
| Missing | 1 (0.2) |  |  |
| **Living environment (%)** |  |  |  |
| Living alone | 308 (65.1) |  |  |
| Living with family | 127 (26.9) |  |  |
| Living with a partner | 20 (4.2) |  |  |
| Living alone: family or relatives nearby | 18 (3.8) |  |  |
| **Household size** |  |  |  |
| Mean | 1.7 | 1.3 | 1.6-1.8 |
| **Comorbidity, no. (%)** |  |  |  |
| None | 401 (84.8) |  |  |
| Asthma | 37 (7.8) |  |  |
| Anxiety disorder | 8 (1.7) |  |  |
| Depression | 6 (1.3) |  |  |
| Hypertension | 5 (1.1) |  |  |

|  |  |
| --- | --- |
| Heart diseases | 3 (0.6) |
| Dyslipidemia | 2 (0.4) |
| Other | 24 (5.1) |
| **Total number of participants** | 473 |

Abbreviations: CI, confidence interval; SD, standard deviation.
<sup>a</sup> Of the 473 respondents, 2 did not specify their age.

### COVID-19 related survey items

**Table 2** and **Supplementary Figure 1** show respondents’ answers to the COVID-19 related survey items. Eighty-one (17.2%) responded that they were either “likely” or “very likely” to contract COVID-19 during the ongoing pandemic; 275 (58.1%) agreed or strongly agreed to a prompt “I feel worried about COVID-19.” Regarding the breakdown of students’ concerns about COVID-19, 182 (38.5%), 121 (25.6%), and 235 (49.7%) acknowledged that they were concerned about the negative impacts of COVID-19 on their future career formation, relationship with teachers, and ongoing research or extracurricular activities, respectively, while 141 (29.8%) also reported concern about a shift toward online education. The reasons for concerns included ineffective educational effect compared with on-site education (*n* = 92, 65.2%), possible resurgence of the COVID-19 outbreak leading to a sudden change in the curriculum (*n* = 74, 52.5%), and decreased clinical exposure (*n* = 92, 65.2%).

**Table 2.**
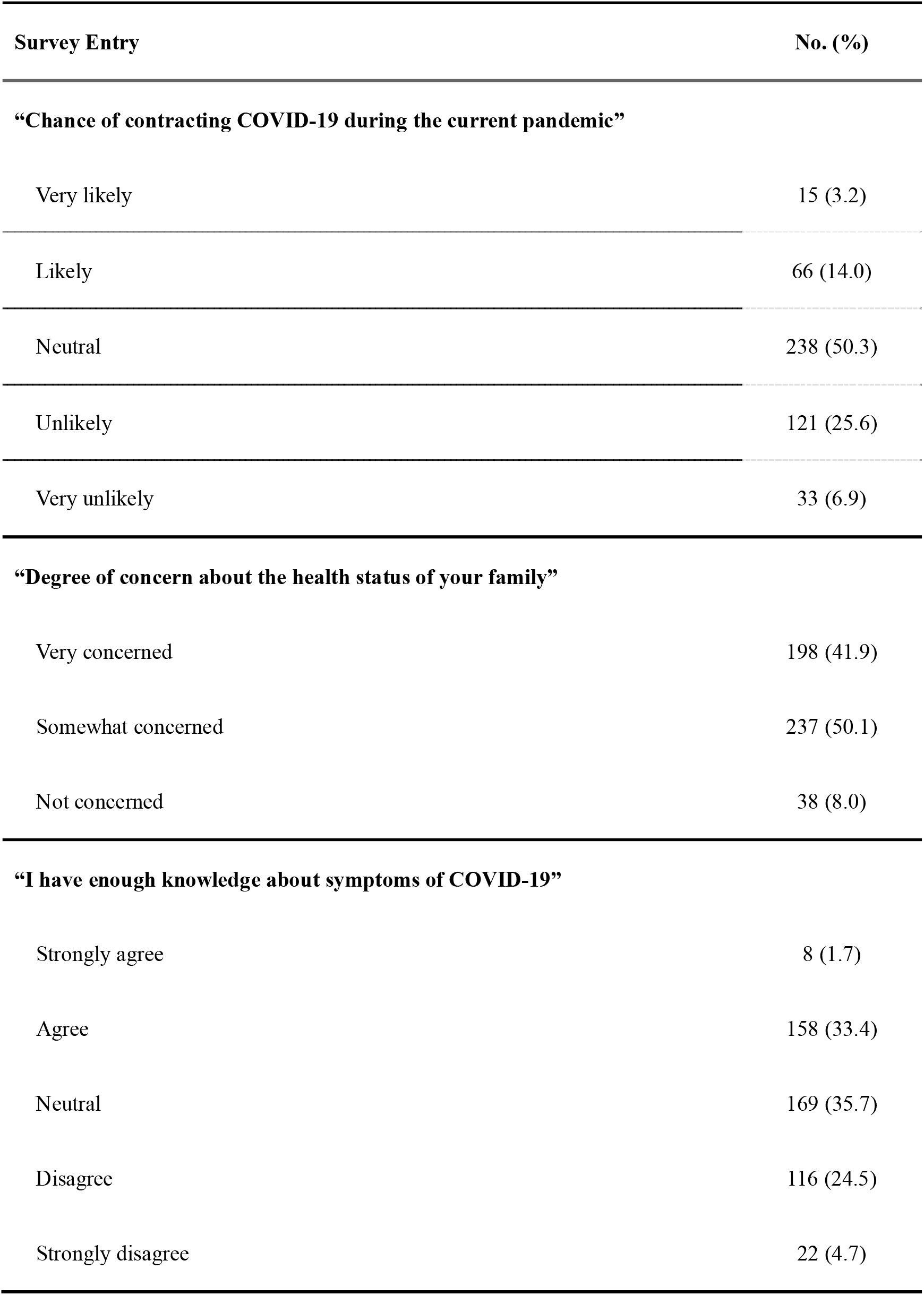
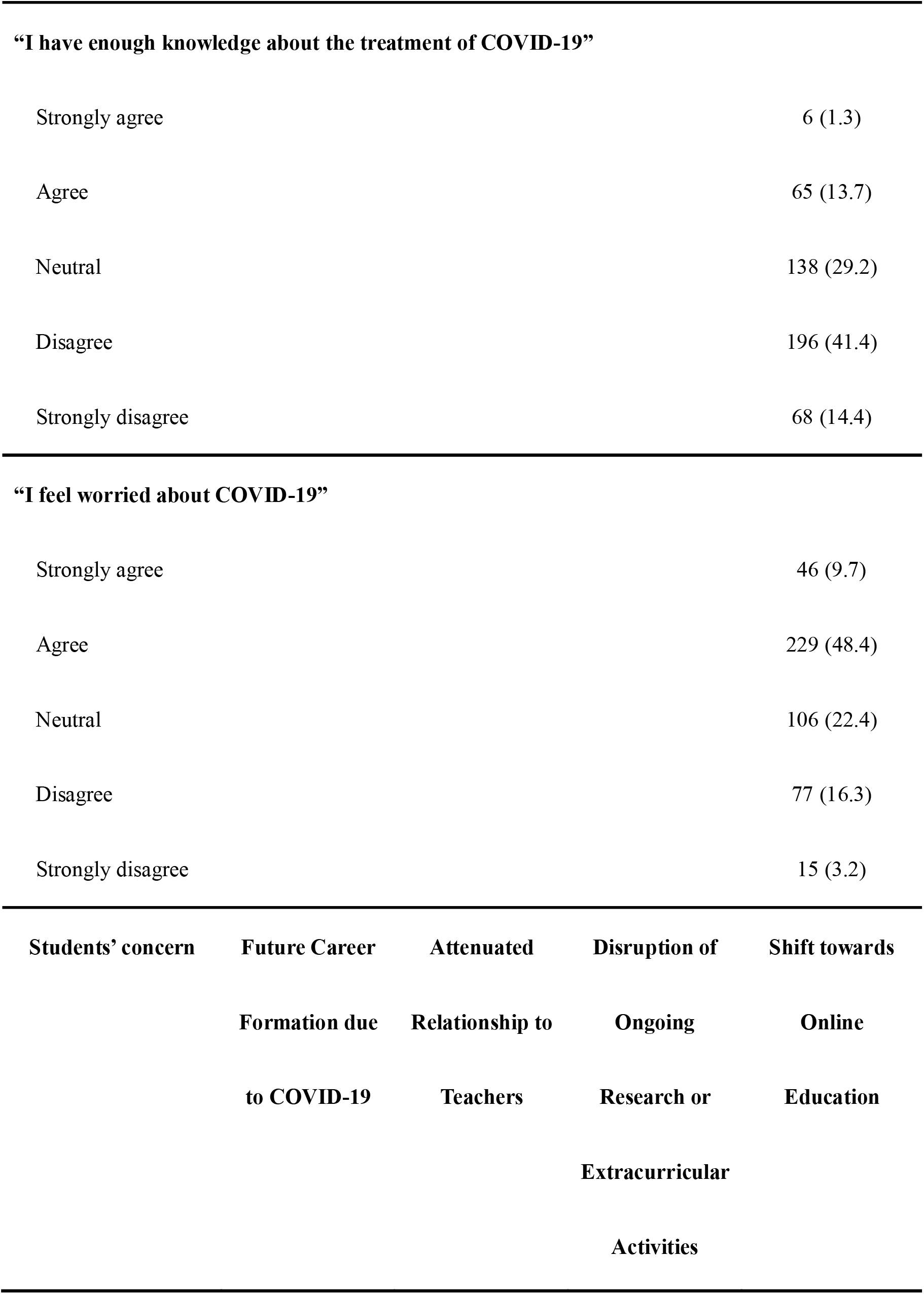
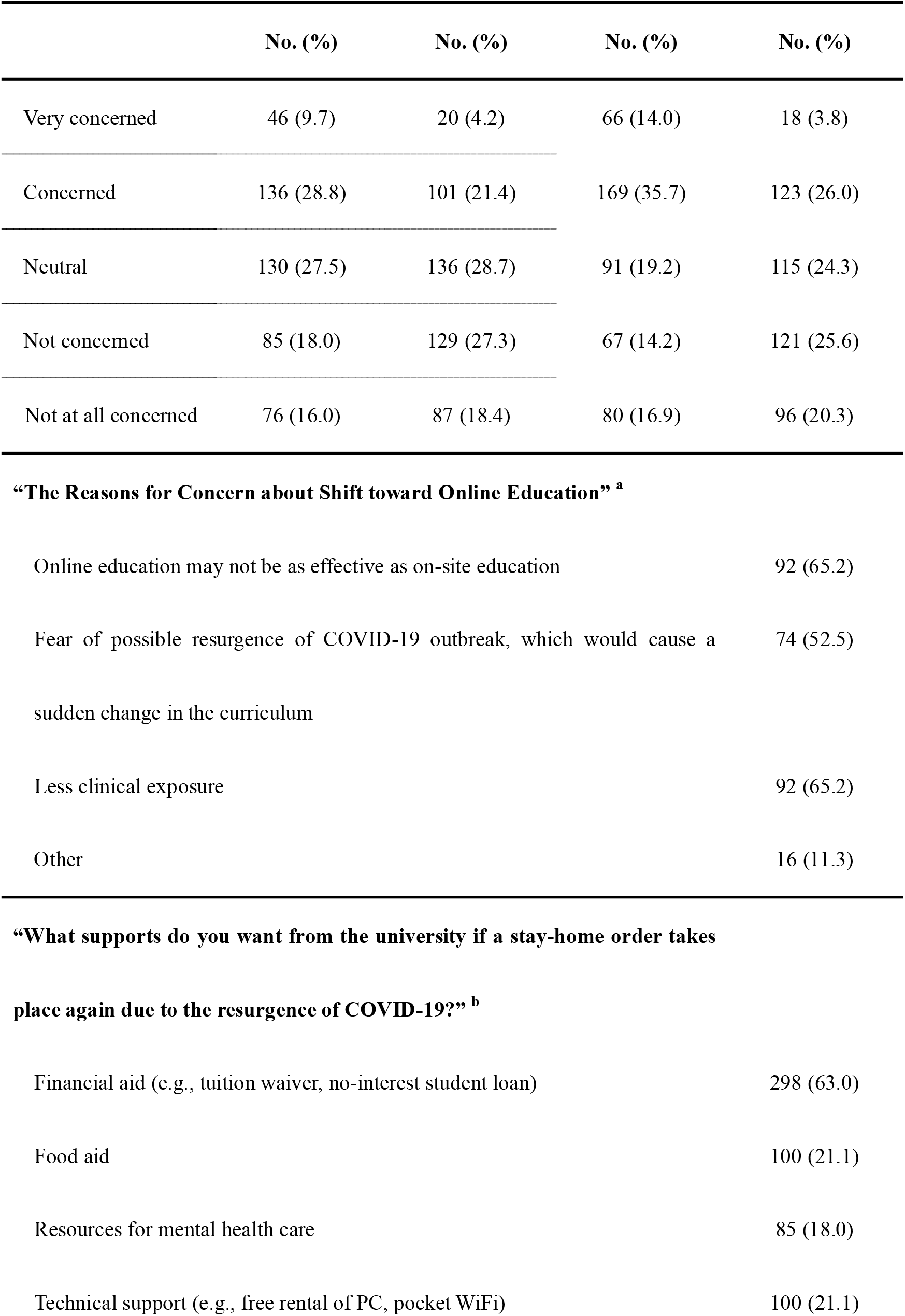
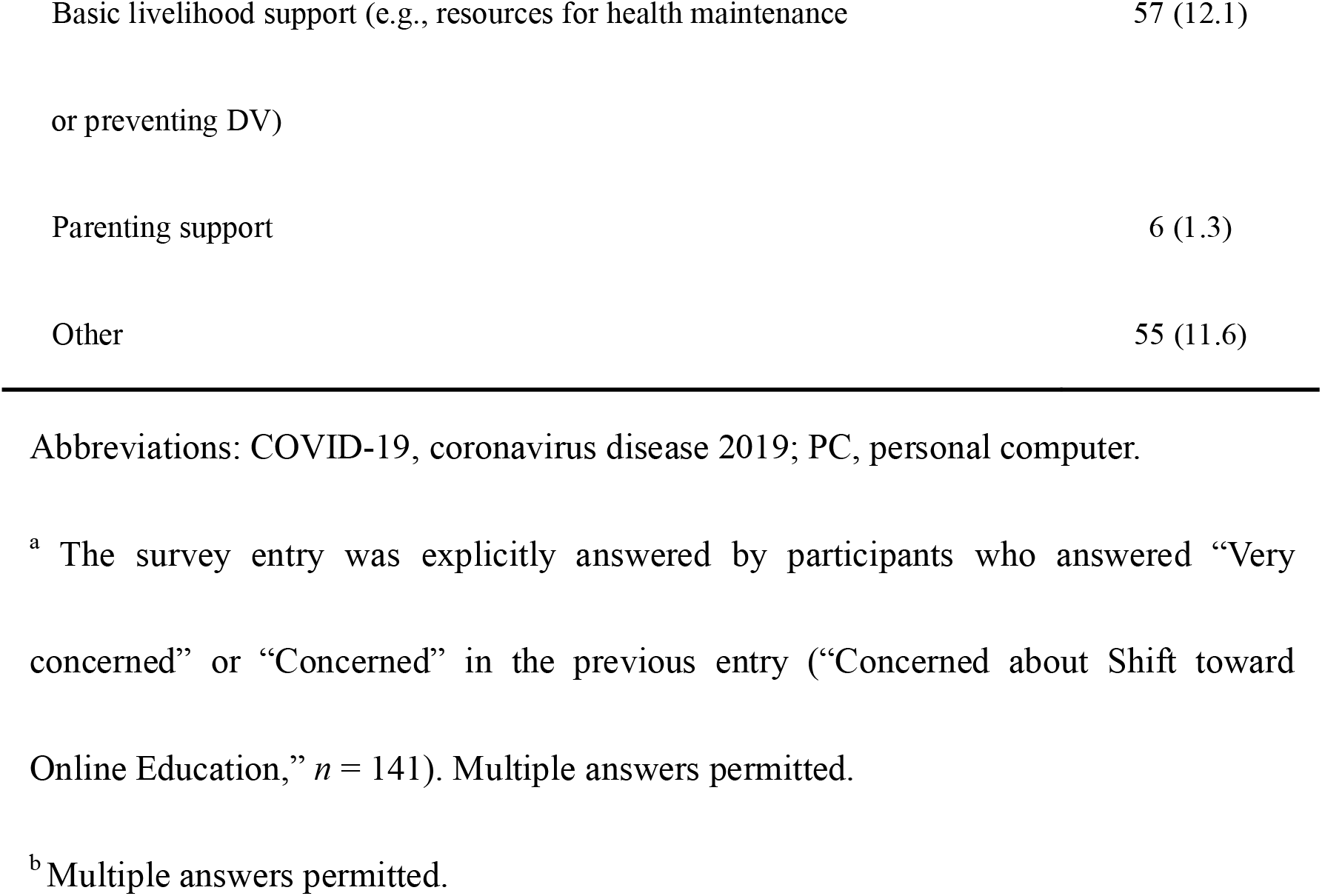
Results of the COVID-19-related Survey Items.

Of the participants, 298 (63.0%) answered that they would request financial aid if a stay-home order recurred due to a COVID-19 resurgence, followed by request for food aid (*n* = 100, 21.1%), technical support for online education (*n* = 100, 21.1%), and mental health care resources (*n* = 85, 18.0%).

### Change in self-learning related time before and after the SOE

We compared the change in average time respondents spent at home, reading books, playing video games, and self-learning per day before the SOE and within two weeks prior to the survey completion (after the SOE). As shown in **Table 3** and **Supplementary Figure 2**, the participants spent significantly longer on all the activities mentioned above after the SOE than before the SOE (*p* <.001). There were also significant differences in their subjective mental health status before and after the SOE based on Wilcoxon signed-rank test *(p* <.001).

**Table 3.** Comparison of Age-weighted Self-learning and Related Activity Time of Medical Students Before and After the National State of Emergency in Japan.

Medical Students Before and After the National State of Emergency in Japan
| Measure | Before SOE |  | Last 2 Weeks<sup>a</sup> |  | *P* value<sup>b</sup> |
| --- | --- | --- | --- | --- | --- |
|  | Time (h) | 95% CI | Time (h) | 95% CI |  |
| Average Time at Home / Day | 16.1 | 15.7-16.6 | 18.8 | 18.5-19.2 | < .001 |
| Average Time Reading Books / Day | 0.70 | 0.58-0.82 | 1.1 | 0.96-1.2 | < .001 |
| Average Time Playing Video Games / Day | 1.5 | 1.3-1.7 | 2.1 | 1.9-2.3 | < .001 |
| Average Time Self-learning / Day | 2.6 | 2.4-2.8 | 4.0 | 3.7-4.2 | < .001 |

| Measure | Before SOE | Last 2 Weeks |
| --- | --- | --- |
|  | No. (%) | No. (%) |
| **Subjective Mental Health Status <sup>c</sup>** |  |  |
| Very good | 85 (18.0) | 39 (8.2) |
| Good | 171 (36.1) | 58 (12.2) |
| Fair | 193 (40.8) | 286 (60.5) |
| Bad | 18 (3.8) | 76 (16.1) |
| Very bad | 6 (1.3) | 14 (3.0) |

Abbreviations: CI, confidence interval; h, hour; SOE, state of emergency.
<sup>a</sup> The base date of the “last 2 weeks” is the day when the participants answered the
survey. The survey period was from June 8 to 14, 2020.
<sup>b</sup> $P$ value is calculated with Wilcoxon signed-rank test.
<sup>c</sup> $P < .001$ with Wilcoxon signed-rank test. There is a significant difference in the subjective mental health status of participants before the Japanese national state of emergency and the last two weeks prior to the date of answering the survey.

### Regression analyses of factors associated with depression and anxiety

Of the participants, 75 (15.8%) had PHQ-9 scores of 10 or more, and 34 (7.2%) had GAD-7 scores of 10 or more. **Table 4** presents all the results of univariate regression analyses. The odds of being depressed were significantly higher in those who had concerns about a shift toward online education (OR 1.97, 95% CI 1.19 – 3.28), and in those who requested food aid (OR 1.99, 95% CI 1.16 – 3.44) and mental health care resources (OR 3.56, 95% CI 2.07 – 6.15) from the university in the event of resurgence of COVID. Regarding generalized anxiety, the odds were higher in respondents who requested food aid (OR 2.50, 95% CI 1.21 – 5.20) and mental health care resources (OR 3.16, 95% CI 1.51 – 6.59).

**Table 4.**
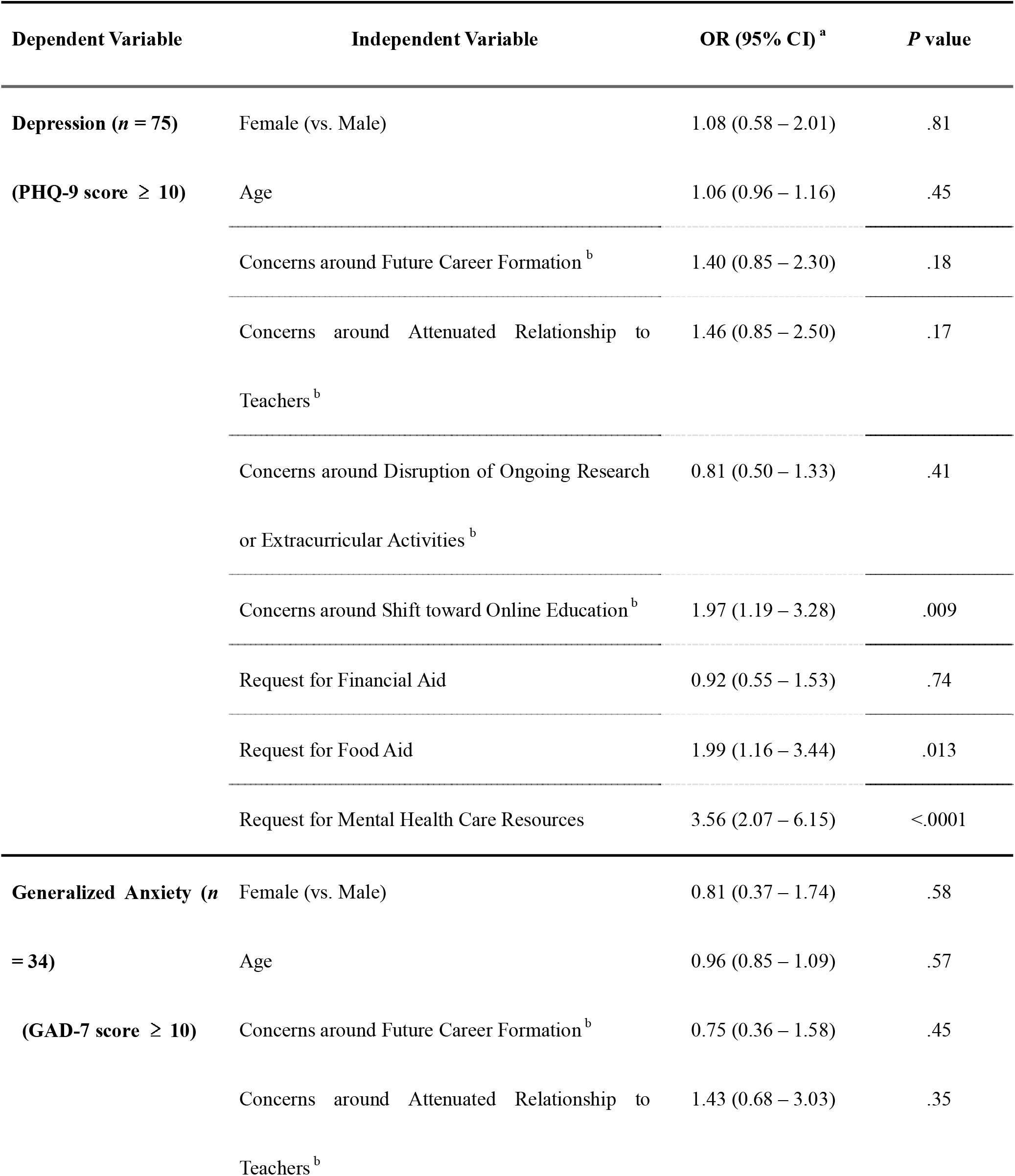
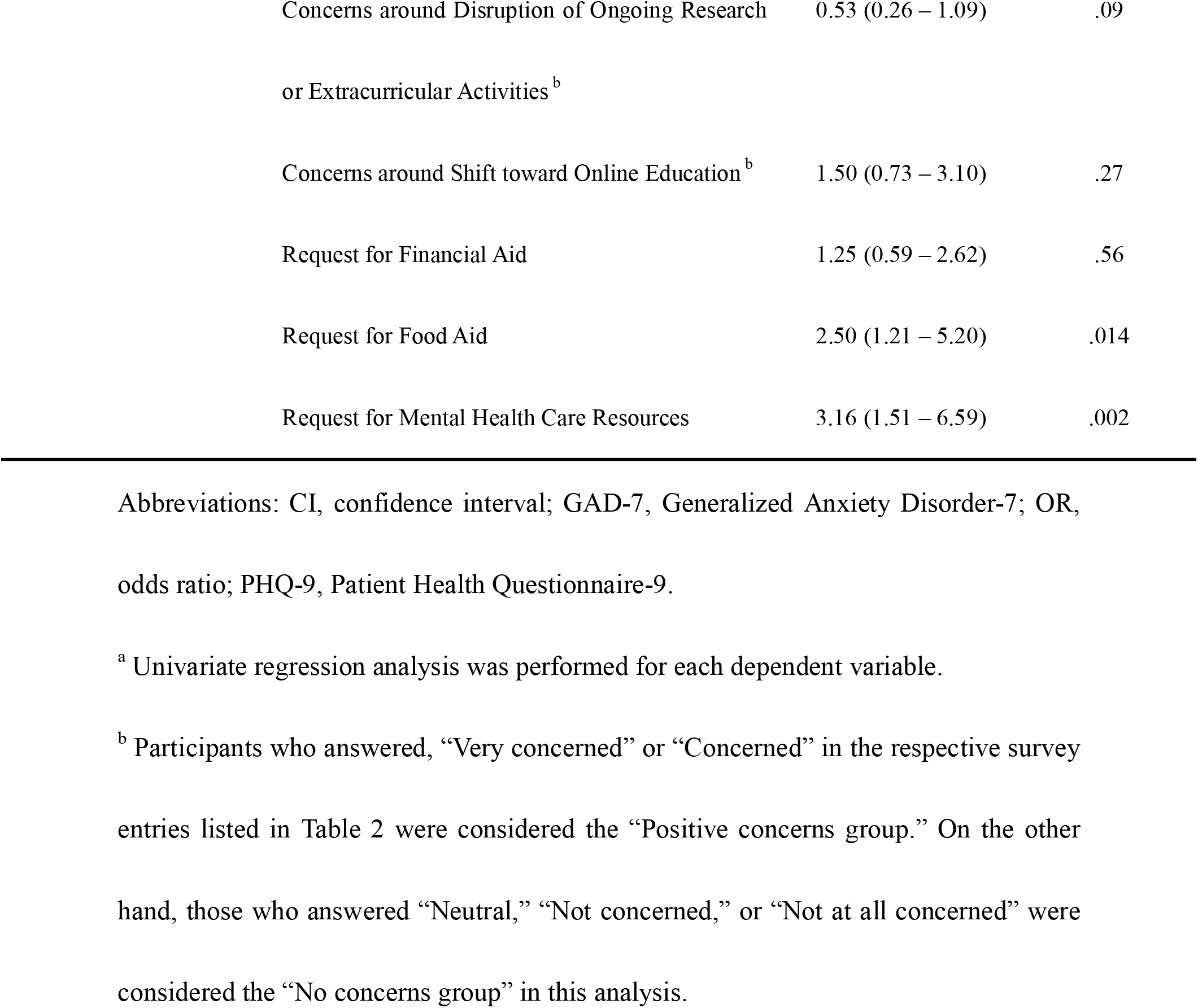
Results of Univariate Regression Analyses of Factors Associated with Depression and Generalized Anxiety in Japanese Medical Students.

### Prevalence of financial hardship

**Table 5** and **Supplementary Figure 3** show the self-reported amounts of monthly allowance from parents and situations regarding financial aid. A total of 332 (70.2%) respondents had a monthly allowance that was lower than the national average (72,810 JPY, approximately 680 USD)(17), while 141 (27.7%) received some sort of financial aid for their living and education.

**Table 5.**
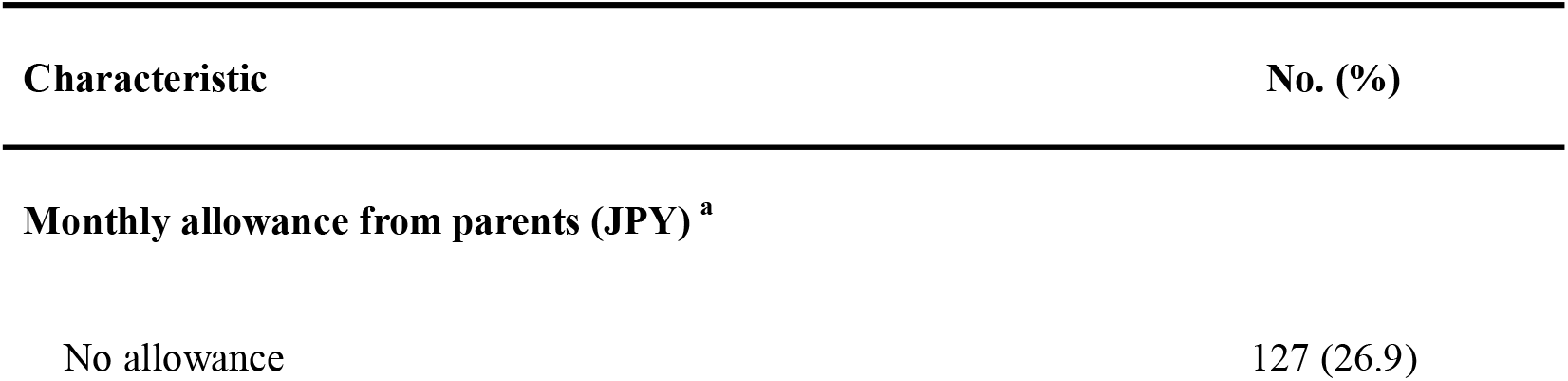
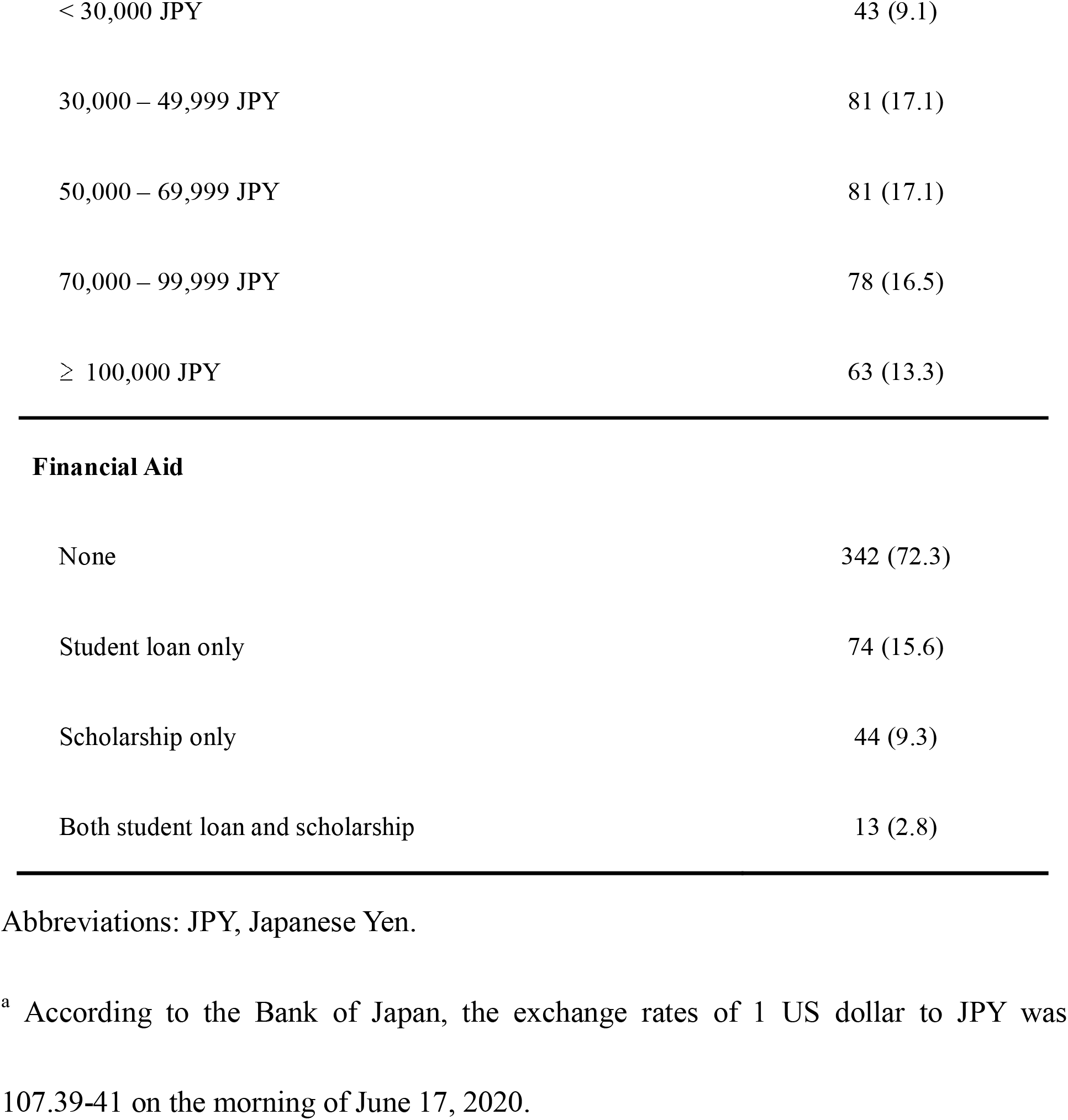
Description of the Financial Situation of Japanese Medical Students.

| Characteristic | No. (%) |
| --- | --- |
| **Monthly allowance from parents (JPY) <sup>a</sup>** |  |
| No allowance | 127 (26.9) |
| < 30,000 JPY | 43 (9.1) |
| 30,000 – 49,999 JPY | 81 (17.1) |
| 50,000 – 69,999 JPY | 81 (17.1) |
| 70,000 – 99,999 JPY | 78 (16.5) |
| ≥ 100,000 JPY | 63 (13.3) |
| **Financial Aid** |  |
| None | 342 (72.3) |
| Student loan only | 74 (15.6) |
| Scholarship only | 44 (9.3) |
| Both student loan and scholarship | 13 (2.8) |

Abbreviations: JPY, Japanese Yen.
<sup>a</sup> According to the Bank of Japan, the exchange rates of 1 US dollar to JPY was 107.39-41 on the morning of June 17, 2020.

## Discussion

To the best of our knowledge, this study is the first survey of Japanese medical students regarding their life circumstances and challenges due to COVID-19. Although Japan has had fewer COVID-19 cases than the US, and Okayama has had comparatively fewer cases than other Japanese prefectures, the study results underscore that the pandemic caused a profound negative socio-educational impact on our society and medical students. We found that approximately 10% of the students on part-time jobs had been fired due to the pandemic. Furthermore, a considerable number of medical students had concerns around their basic life security, demanding support from the university in the form of financial aid, food aid, technical support, and mental health care resources due to the SOE. Regarding students’ subjective psychological distress, those who expressed concerns about the rapid shift toward online education and fear around basic life security were more likely to be depressed and anxious after the SOE.

Concerns around future career disruption, attenuated relationship to medical teachers, and disruption of ongoing extracurricular activities were prevalent among the participants, which underlines the considerable uncertainty amid the COVID-19 pandemic (Table 2). In particular, 63% of respondents reported the need for financial aid in the event of a second wave of the pandemic. These data correspond to the fact that more than 70% of participants received no more than the national average monthly allowance from their parents (Table 5). Contrary to the general notion in Japan that “medical students are financially well-off,” many may have experienced financial hardship. Recently, the theory of willpower has gathered public attention. Financial stability is considered a prerequisite of willpower, which aids appropriate decision-making(18). Combined with previous research findings that financial instability could lead to worse mental health outcome(19, 20), educators are expected to develop a strategy to offer financial support to those in need.

Regarding lifestyle changes and psychological distress, after the SOE was lifted, the participants experienced significantly worse mental health status, and spent significantly longer at home, reading books, playing video games, and learning by themselves (Table 3) than before the announcement of SOE. Our results are consistent with prior longitudinal and cross-sectional studies that the COVID-19 pandemic led to a sedentary life and psychological distress(9, 12, 13, 21). As suggested by a previous study, the increase in gaming could be a coping behavior against psychological stress(22).

As for regression analyses of factors associated with depression and anxiety, in our study population, those who requested food aid and mental health care resources from the university upon the future resurgence of the COVID-19 outbreak had significantly higher odds of depression and anxiety. Furthermore, concerns around the shift toward online education were identified as a factor associated with depression (Table 4). Surprisingly, 65.2% of those who had concerns about the shift toward online education thought online education was less effective than in-person education. While previous studies have reported the utility and noninferiority of online learning compared to offline(23, 24), in-person learning, the results of the current study revealed a potential gap in perception regarding the effectiveness of online education between medical students and educators. Educators should not assume that students know the potential benefits of online learning, and it is essential to inform learners that online learning is non-inferior to in-person learning, which could attenuate potential depression and anxiety. While in-person communication has become difficult nowadays due to the fear for COVID-19, medical schools may need to reach out to students to find out those suffering from underlying life insecurity and to provide multilateral support, including early mental health care interventions.

Several limitations to this study should be noted. First, due to the single-center cross-sectional survey design, we may not conclude causal relationships. Second, the cross-sectional research contains a limitation in terms of addressing changes over time. While we illustrated the changes in students’ average time spent on self-learning related activities before and after the SOE in Table 3, a longitudinal study design would be more appropriate to examine the differences in the study cohort. Third, we asked participants to provide their mental health status and time spent on self-learning related activities before the SOE (approximately six weeks before the survey implementation), both of which are subject to recall bias. Also, PHQ-9 and GAD-7 scores were obtained only after the SOE. Thus, it is possible that they might have not changed during the period. Lastly, due to the nature of the survey topic, those who were interested in the public health emergency or mental health may have been more likely to respond, which would lead to self-selection bias. Despite these limitations, a total population sampling strategy coupled with higher-than-usual response rates(25) contributed to high internal validity.

In conclusion, through the study, we have provided graphical data and evidence regarding the socio-educational impacts of the COVID-19 pandemic on medical students. In circumstances of considerable uncertainty, both educators and medical students need to be flexible, patient, and resilient. The uncertainty and drastic change triggered substantial psychological distress in students, which was greater than we assumed. As discussed, Japan has had a relatively small number of COVID-19 cases compared to the US and other European countries. Although Okayama prefecture survived the pandemic with fewer confirmed COVID-19 cases than other prefectures, medical students experienced significant impacts due to the public health emergency. The educational institution should recognize the prevalence of basic needs insecurity, such as financial difficulties and a shortage of staples, including food, etc. As medical educators, we need to be accountable for the advantages of online education in the field of medicine to alleviate students’ psychological distress, in addition to providing multilateral support to those in need, including early mental health care interventions. While we targeted medical students in a single Japanese national university, the survey results warrant further research and analysis if the distress is amplified by existing anxiety, depression, burnout, etc., or is a wholly COVID-19-related phenomenon. We call for increased research in populations with more COVID-19 cases than Japan, to figure out the challenges medical students have been facing amid the pandemic in different cultures and backgrounds.

## Acknowledgments

We would like to thank the Academic Affairs Division of Okayama University Medical School for its cooperation in conducting the survey.

## Author contributions

YN wrote the manuscript, designed the study, and analysed the data. KO, KT, MO, HH, and HK analyzed the data and revised the manuscript. FO designed and supervised the research.

## Figure Legend

**Supplementary Figure 1**

(A) The breakdown of students’ concerns about COVID-19. 182 (38.5%), 121 (25.6%), and 235 (49.7%) acknowledged that they were concerned about the negative impacts of COVID-19 on their future career formation, relationship with teachers, and ongoing research or extracurricular activities, respectively, while 141 (29.8%) also reported concern about a shift toward online education. (B) Of the participants, 298 (63.0%) answered that they would request financial aid if a stay-home order recurred due to a COVID-19 resurgence, followed by request for food aid (n = 100, 21.1%), technical support for online education (n = 100, 21.1%), and mental health care resources (n = 85, 18.0%).

**Supplementary Figure 2**

The bar charts show the change in average time respondents spent at home, reading books, playing video games, and self-learning per day before the SOE and within two weeks prior to the survey completion (after the SOE). Error bars show the 95% confidence intervals.

**Supplementary Figure 3**

The self-reported amounts of monthly allowance from parents and situations regarding financial aid are shown.

