## Supplementary figures and images for "Socio-educational Impact and Psychological Distress of Medical Students amid the COVID-19 Pandemic: A Japanese Cross-Sectional Survey"

### Supplemental Figure 1

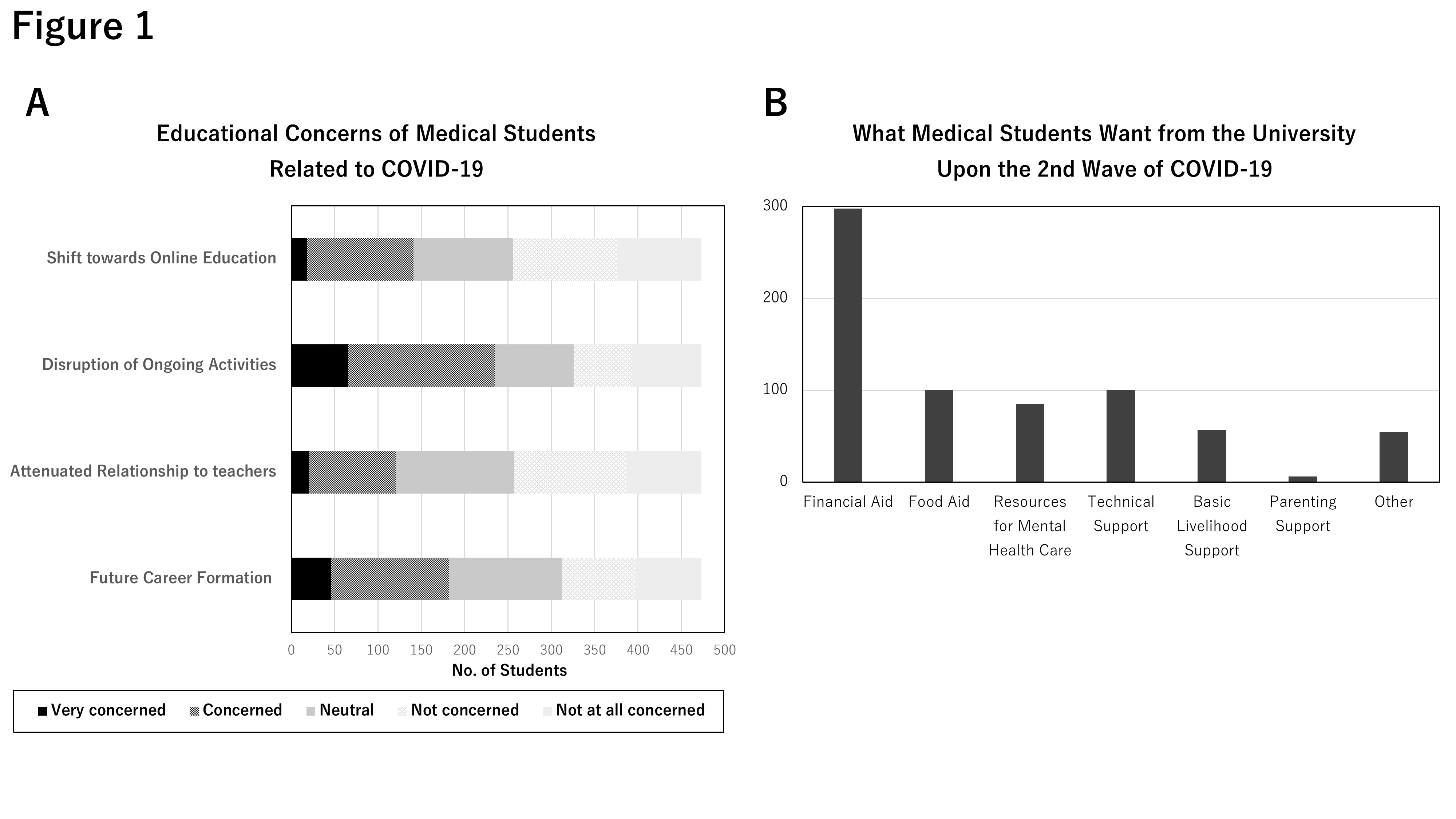

### Supplemental Figure 2

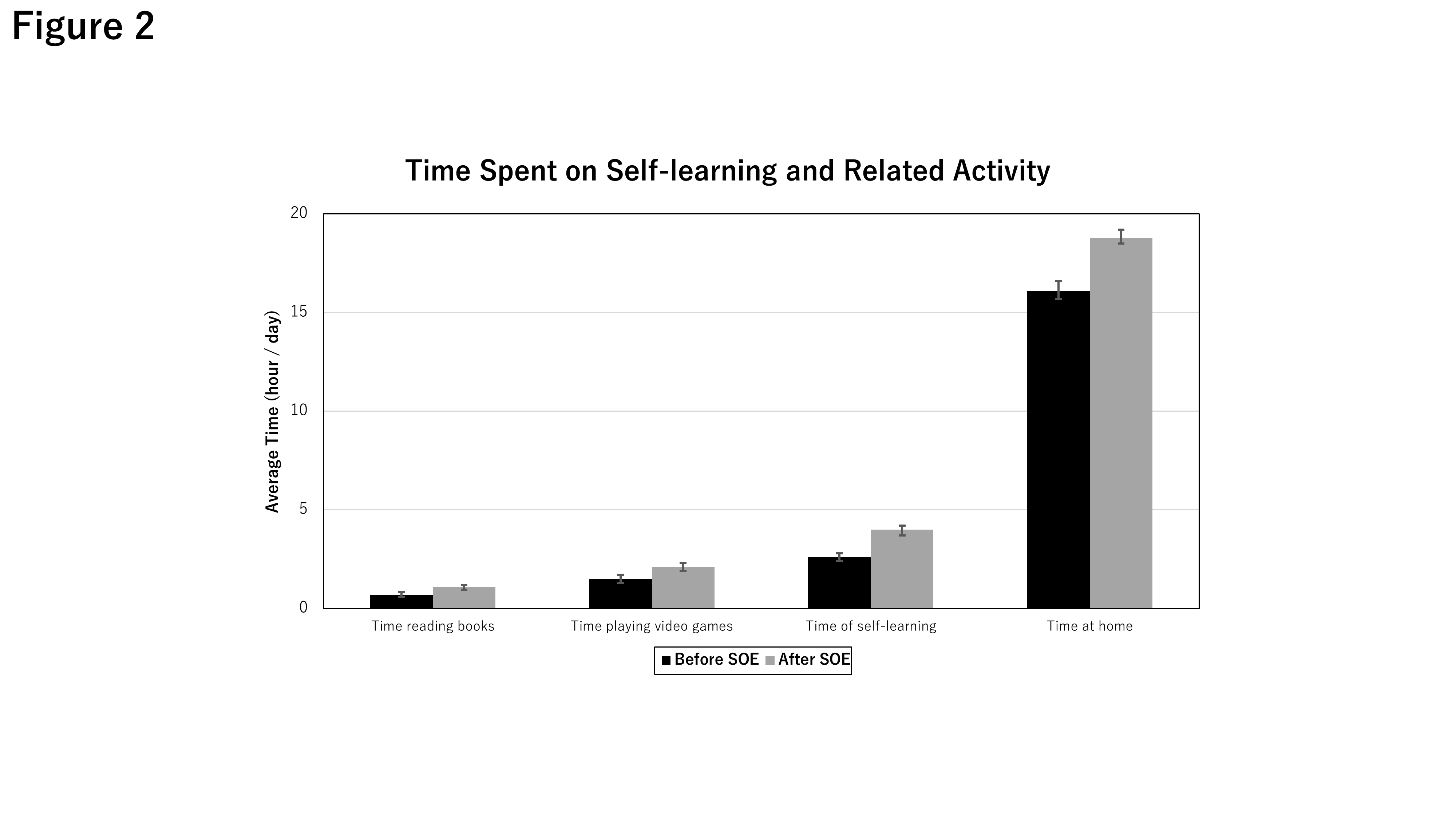

### Supplemental Figure 3

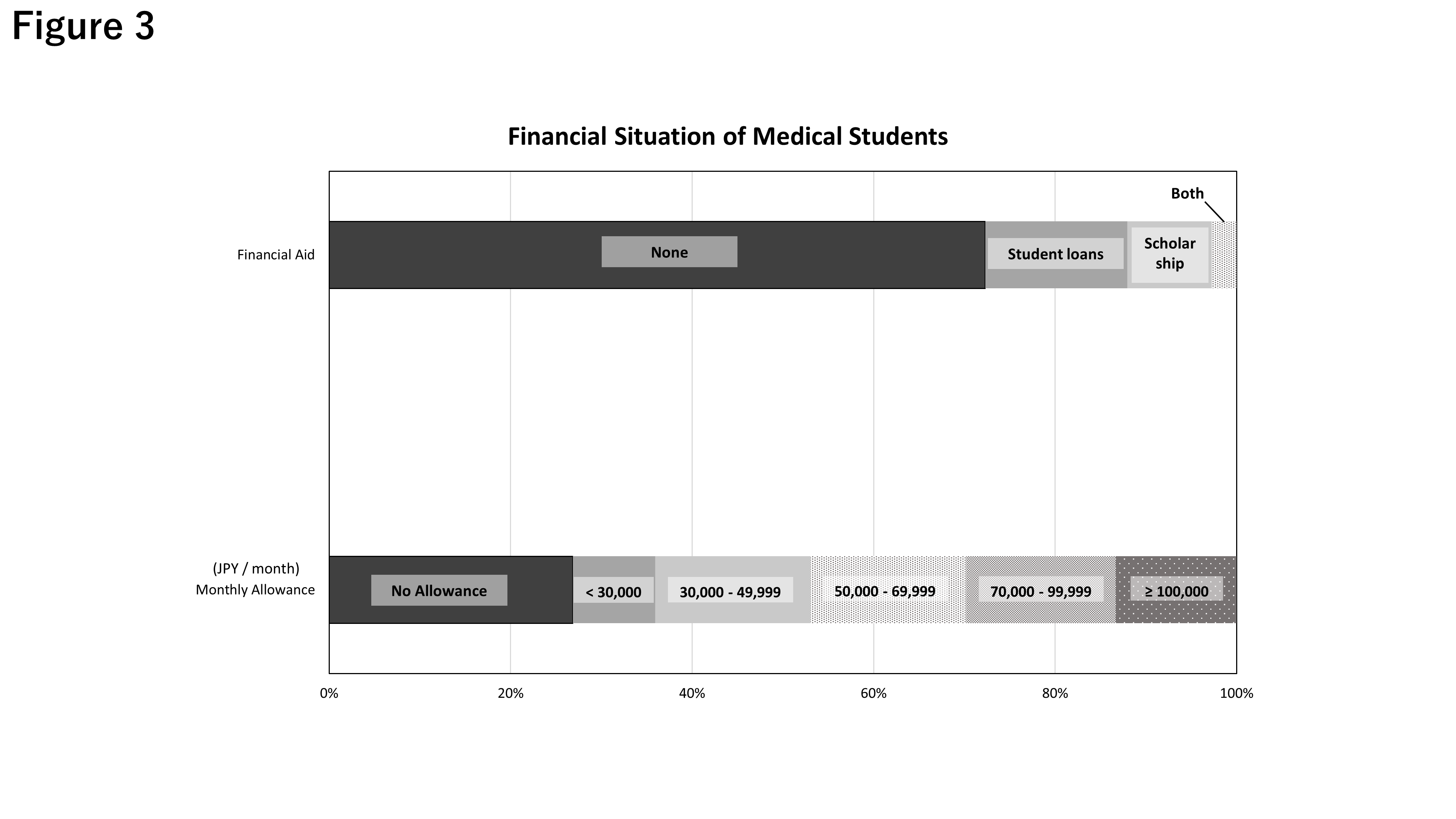
